# Modeling projections for COVID-19 pandemic by combining epidemiological, statistical, and neural network approaches

**DOI:** 10.1101/2020.04.17.20059535

**Authors:** Steffen Uhlig, Kapil Nichani, Carsten Uhlig, Kirsten Simon

## Abstract

As the number of people affected by COVID-19 disease caused by the novel coronavirus SARS-CoV-2 ebbs and flows in different national and sub-national regions across the world, it is evident that our lifestyle and socio-economic trajectories will have to be adapted and adjusted to the changing scenarios. Novel forecasting tools and frameworks provide an arguable advantage to facilitate this adapting and adjusting process, by promoting efficient resource management at individual and institutional levels. Based on deterministic compartment models we propose an empirical top-down modeling approach to provide epidemic forecasts and risk calculations for (local) outbreaks. We use neural networks to develop leading indicators based on available data for different regions. These indicators are not only used to assess the risk of a (new) outbreak or to determine the effectiveness of a measure at an early stage, but also in parametric models to determine an effective forecast, along with the associated uncertainty. Based on initial results, we show the performance of such an approach and its robustness against inherent disturbances in epidemiological surveillance data. We foresee such a statistical framework to drive web-based automatic platforms to democratize the dissemination of prognosis results.

## INTRODUCTION

How can the course of the pandemic be predicted, what can be expected in the short term, when can the peak number of infections be expected, when can the number of new infections decrease to a manageable level? Answers to such questions are sought after by one and many. But forecasting and predictions during a crisis such as the one we are in, is a double-edged sword^1,2^. As the adage goes, that a model is only as good as the assumptions and the data used to ‘train’ one; the risk of incorrect predictions or unreliably large uncertainty intervals is fatal. This is the reason there is a high level of skepticism over any and every forecasting model proposed in recent week(s). Consider weather forecasts in our daily lives. These are so ubiquitous that, on the one hand, we often fail to acknowledge the strides made in meteorological modeling^3^. On the other, we tacitly recognize without frowning that predictions can go astray. Unlike weather forecasts, epidemic (or pandemic) projections are highly complex and challenging. Nevertheless, having reliable estimates for morbidity and mortality is critical for decision making at individual and institutional levels^4^. As different parts of the world prepare to return to a new normal, in which there is a continued risk of rapid spread as soon as physical distancing measures are reduced^5^; there is a need for a new set of approaches targeted specifically at sensing and predicting the progression of the disease. Through a novel modeling approach rooted in multi-modal data, the aim here is to characterize the threats and impacts for assessing effects of interventions and facilitate decision making.

### Uncertainty of model parameters

Compartmental models such as SIR and SEIR (and their variants) are necessary to better understand the mechanisms that come into play through the course of an epidemic^6^. However, the suitability of these models to make prognosis remains to be exploited. From a statistical perspective, multiple constraints and challenges exist in realworld applications of epidemiological “bottom-up” models to make a meaningful prediction. Predominantly, the availability of surveillance data on different compartments such as susceptible, exposed, infected, or removed (recovered) is limited. In view of the large disparity in testing capacities, and lack of clarity on unreported cases, the models derived thereof cannot provide an unequivocal picture of the epidemic potential. Adding to these challenges is the highly fluctuating nature of daily reported figures. The uncertainties associated with each of these factors, snowball to a monumentally large uncertainty value for the outcome of the built model^7^. Besides, the predictions derived from the models can become unstable because the figures are often susceptible to small changes in the input parameters. Estimation procedures are needed to specify the model parameters, which in turn rely on the existing, very noisy time series data. This not only applies to the simplest SIR model, but also models based on individualized assumptions for different subpopulations. In order to use such models for forecasting, one needs reasonable assumptions for each individual model parameter. Notwithstanding the fact that increasing number of model parameters increases the scope of possible errors for the respective forecast. As an example, the revision of estimated number of deaths due to COVID-19, in subsequent reports, by the expert team at Imperial College, London, sparked several reactions^8^. The models initially projected around 500,000 deaths to occur in the UK, alone. The number was updated to under 20,000 deaths. This happened just in one to two weeks and demonstrated that any prognoses in an ongoing epidemic may fail^9,10^. One of the scenarios that was also reported as a possibility (at the time of publication) that, half of the population was already infected by SARS CoV-2. This sounds extremely unlikely, given the fact that about three weeks later, the numbers barely crossed 100,000 confirmed infected cases. Nevertheless, during this initial phase, it was advisable to consider even such extreme scenarios due to the very contradictory information situation. Similarly, commentaries and responses have been spawning in reaction to the (daily) changing estimates of the model^11^ developed by the IHME team at University of Washington^12^.

### Theoretical bottom-up versus empirical top-down modeling

Building over the widely adopted bottom-up approaches (based on SIR, SEIR, SIA, and their variants), we propose in this work an empirical top-down approach. For such an approach, the starting point is not the epidemiological description of the disease with all its individual aspects (bottom-up), but instead, the resulting data with all its errors (top-down). The method does not claim to represent a simulation of the reality. Still, it aims to derive underlying trends and (suitable) risk indicators from the existing noisy time series data for confirmed infected cases and deaths. These indicators are derived both from epidemiological principles and pertinent statistical theory, with special focus on systematic and random errors. The indicators selected by means of neural network analysis not only serve as early warning indicators, but also form the basis of the statistical models used for the forecast. Hence, these indicators have a two-fold role – (a) assist in evaluating the ‘current’ epidemic potential and (b) enable reliable forecasts.

In the following sections, we outline the adopted methodology, provide preliminary findings at a country level (while arguing that going down to a higher level of granularity is important), and relevant implications of the findings. We demonstrate how the approach has been integrated into a web-based portal enabling automatic and real-time evaluation of metrics, with quantified uncertainty to provide forecasts for districts and cities of Germany. These results can equip decision-makers to enable primary containment or mitigation strategies.

## METHODS

Because the evolution of the pandemic is extremely dynamic, classical statistical procedures and methodologies for forecasting, in our view, are inadequate for making viable projections. Trends are highly non-linear, and extrapolating the case counts, whether linear or exponential, does not usually lead to the desired outcome^13^. Therefore, the method adopted here involves fusing mechanistic and statistical approaches. Consider time series data for the number of daily new confirmed cases, and the number of daily deaths reported. Given, for instance, the time-series data has fifty time points, thousands of different potential indicators can be derived by a statistician familiar with time-series analyses. For example, by calculating successive differences of the first and second order. Additionally, averages of consecutive numbers, along with simple relative change calculations can be made. Furthermore, it makes sense to use different data sources in parallel, to improve the forecast on this basis. To evaluate which indicators are appropriate and wield a higher prognostic value, are usually checked manually using proven methods of time series analysis. In order to expedite this very lengthy process, one can also make use of artificial intelligence (neural network) methods. This is exactly the method we use for the forecasting framework of COVID-19. The timeseries data available at a certain point in time form the basis for the training of the neural networks. The corresponding short and medium-term forecast values are used as the labels for the training. Feature selection is then used to extract significant metrics. An appropriate parametric model is developed using these metrics as covariates. Such a parametric model then allows assigning a quantitative context to relevant factors along with its uncertainty. Based on the neural networks, we obtain not only one, but several promising indicators. These are then fed, into competing parametric models, to not only obtain a forecast that is as accurate as possible, but also to get an idea of the range of possible outcomes when using different data sources and different tools for prognoses.

As we are currently in the middle of a rapidly progressing pandemic, many forecasts have a short time horizon of a few days, weeks, or at best months. The basis of the forecasts is usually the number of confirmed infections or the number of deaths. However, the quality of the reported figures, i.e., in particular, whether the information is correct, whether there are random and systematic deviations and how reliably and quickly they are reported, can be subject to strong fluctuations from country to country and from region to region. Such wavering data flows also show variations over time. For example, the number of confirmed infections is not a particularly suitable measure of the current spread of the epidemic if there is a lack of adequately coordinated testing strategy in the different regions, i.e., if the selection criteria for the persons to be tested are not the same. It becomes even less favorable if the selection criteria change over time or if the criteria for a confirmed diagnosis changes over time. The latter happened both in China and Germany. As a result, an enormous jump in the cases on 12.02.2020 was observed in China. Moreover, the number of deaths is not always a particularly suitable criterion either; if one considers, for example, that the beginning of the epidemic in Bavaria, Germany, was marked by many young winter holidaymakers in Austria and Italy who were infected but had a very low mortality rate. Only weeks later, when the epidemic increasingly reached older cohorts, did the death rates increase. Thus, if the spread of the epidemic had been modeled on the basis of death rates, only a minimal number of deaths would have been seen at first, but then the number of deaths increased quickly. It would hardly have been possible to deduce the actual growth of the epidemic from these figures. Even if the epidemic has reached all population groups equally, it is not possible to derive any reliable information from the death rates regarding the spread of the virus, especially not when the health care systems reach the limits of their capacity. However, the death rates become relevant when they are considered in combination with other parameters, such as infection rates, or daily count of infected persons based on the day for the onset of symptoms.

Various countries and regions have different socioeconomic structures, demographic distributions, healthcare systems, political will, and culture. Thereby leading to variations in what is reported and what is known. A typical example in the scope of work presented here is the ‘weekend effect’ in reported numbers of confirmed cases across Germany. Many regional authorities do not report numbers over the weekend; consequently, relative lower numbers are consistently reported for the weekend. In our approach, by utilizing a large set of warning indicators, such region-specific effects are rationalized in the model building process. Therefore, depending on the time and situation, a completely different approach to the prognosis of the epidemic may be possible. Additionally, it is also necessary to learn from the experience of those countries that have already survived the first wave of the epidemic, and this, in turn, means that we are dealing simultaneously with very different data and models, each with the same objective, namely to recognize when the wave rises, when it reaches its peak, when it has dwindled again and how many smaller local waves may occur as a result.

## FINDINGS & INTERPRETATIONS

Our preliminary work is based on data sourced from the Center for Systems Science and Engineering (CSSE) at Johns Hopkins University^14^ and other aggregator websites^15^. Until now, we use time-series data for the provinces of China, South Korea, Japan, Germany, Italy, Spain, the UK, the US, and Russia, but as the pandemic is progressing, we are planning to include further data. We identify several candidates for early warning indicators (EWIs) through the neural network approach described above. Most of these EWIs describe the dynamics of the process or compare the dynamic relationship between deaths and confirmed infections.

### Early warning indicators (EWIs)

EWIs have a 2-fold role – (a) evaluate the reliability of the prognostic assessment; in other words, to assess if a reliable prognosis is possible or not, (b) utilize the underlying trends of such indicators in the parametric modeling process. In the following sections, we show the interpretation of one such EWI candidate, also referred to as EWI candidate A as an example for South Korea, Germany and the USA.

### South Korea

Here is an example for an indicator describing the dynamics in South Korea. Values for EWI above 0 indicate an increase in the number of infected persons (reproduction rate R>1), and values for EQI below 0 represent a decrease (reproduction rate R<1). The higher the value, the faster the rate of growing epidemic due to higher transmissibility.

Looking at the course of the indicator for South Korea (Fig. 1A), together with the trend of confirmed infections (Fig. 1B), 5 phases can be distinguished (see Table 1). The trend indicator is suitable on the one hand for defining and understanding the different stages in the course of the epidemic, but also as an early indicator for a (local) outbreak of the epidemic or for the effectiveness of measures to contain it.

**Table 1:** Descriptions for how the early warning indicator, candidate A for South Korea.

| Date range (2020) | Key takeaways |
| --- | --- |
| Feb 1 to Feb 19 | Random fluctuations around 0 (single cases only) |
| Feb 20 to Feb 26 | Outbreak starting in Daegu |
| Feb 27 to Mar 10 | The containment measures gradually slowed down the epidemic |
| Mar 13 to Mar 22 | A break in the trend indicator around 13.03.2020 suggests a renewed outbreak of the epidemic and a second, albeit much smaller wave, which, however, soon collapsed again |
| Apr 1 to Apr 12 | Another decrease of trend indicator since April 1<sup>st</sup> suggests successful containment of the second wave |

**Figure 1:**
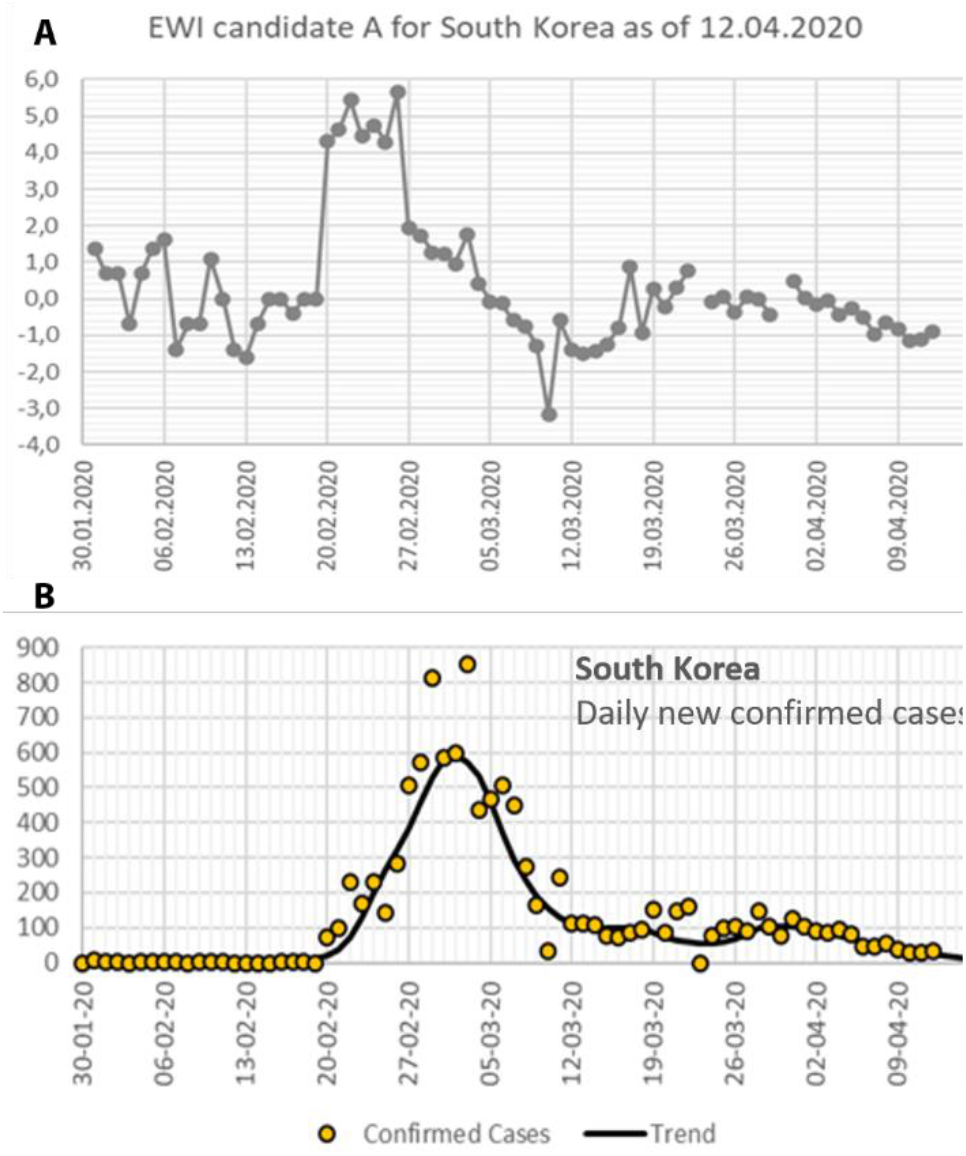
(A) EWI candidate A for South Korea 30.01.2020 – 12.04.2020. EWI candidate is a measure of whether the spread of the epidemic is currently accelerating (>0) or slowing down (<0). (B) Daily confirmed cases in South Korea (Data source: Johns Hopkins University)

Early indicator for an outbreak gives a signal on the 20th of February 2020. From 27th February to 9th March, there is a clear linear downward trend, reflecting that containment measures were effective and finally reduced the number of daily infections; break in the downward trend of the indicator around 11.03.2020 and 14.03.2020 suggests the beginning of another (local) outbreak. Another decrease of trend indicator since April 1st suggests successful containment of the second wave.

### Germany

The same indicator for Germany looks different (see Fig. 2A-B). It appears that Germany has been in a phase of slowing down the epidemic for at least five weeks and is now experiencing a decline in the number of infections.

**Figure 2:**
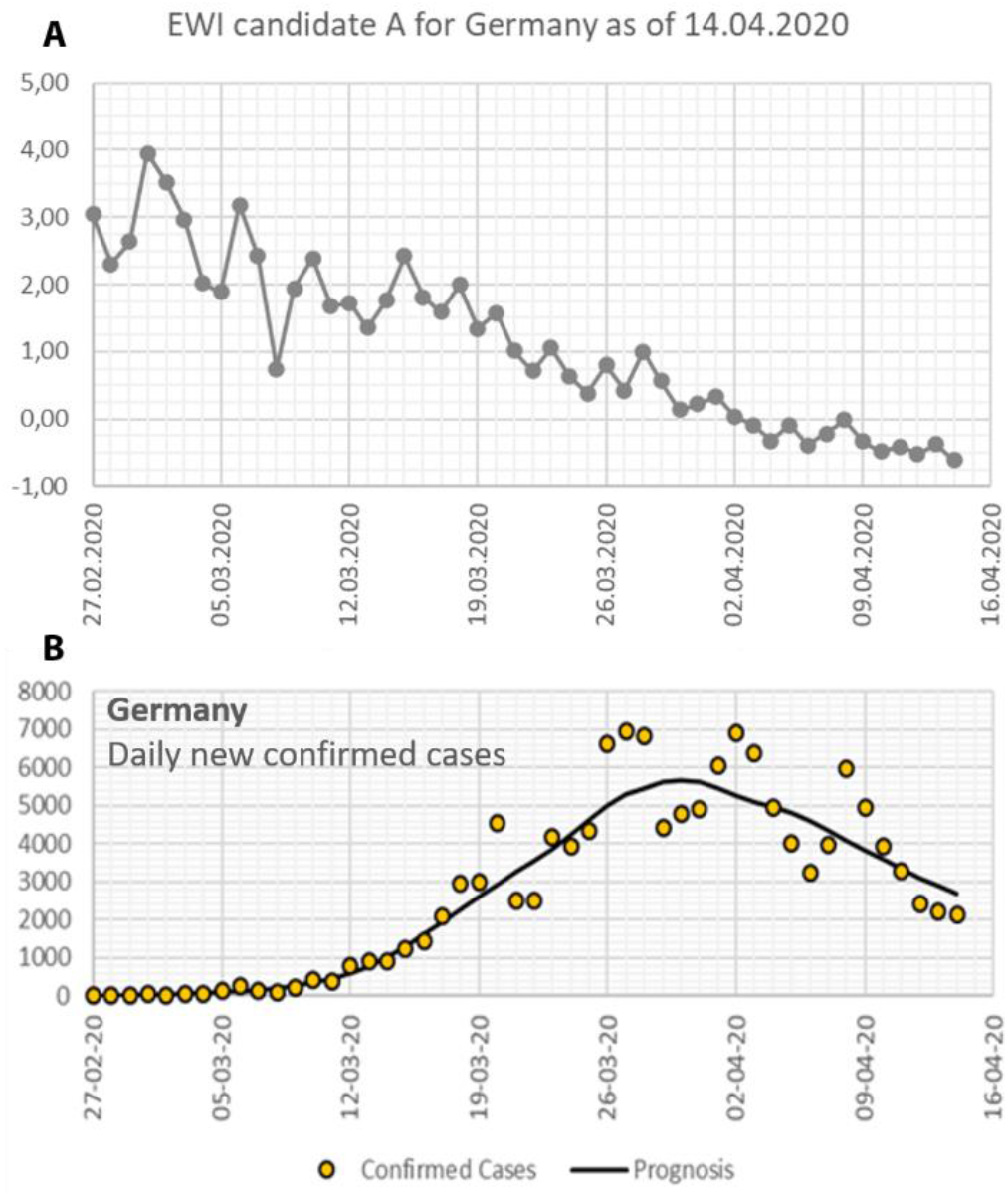
(A) EWI candidate A for Germany 27.02.2020 – 14.04.2020. EWI candidate is a measure of whether the spread of the epidemic is currently accelerating (>0) or slowing down (<0). (B) Daily confirmed cases in Germany (Data source: Johns Hopkins University)

There are no apparent breaks in the overall course; however, erratic fluctuations in infection numbers from day to day make it challenging to identify such effects.

### United States of America

At a first glance, the trend for the same EWI for the US looks like that for Germany. As in the case of Germany, there is a clear decline over the entire period and thus a considerable slowdown in the epidemic. The daily fluctuations for US (see Fig. 3A-B) are considerably smaller than in Germany and reveal that the course of events by no means follows a simple linear trend. Rather, it shows that this trend has weakened considerably over the three weeks. Since the indicator is currently still fluctuating around the value 0, the further decline is not yet very well supported from a statistical perspective.

**Figure 3:**
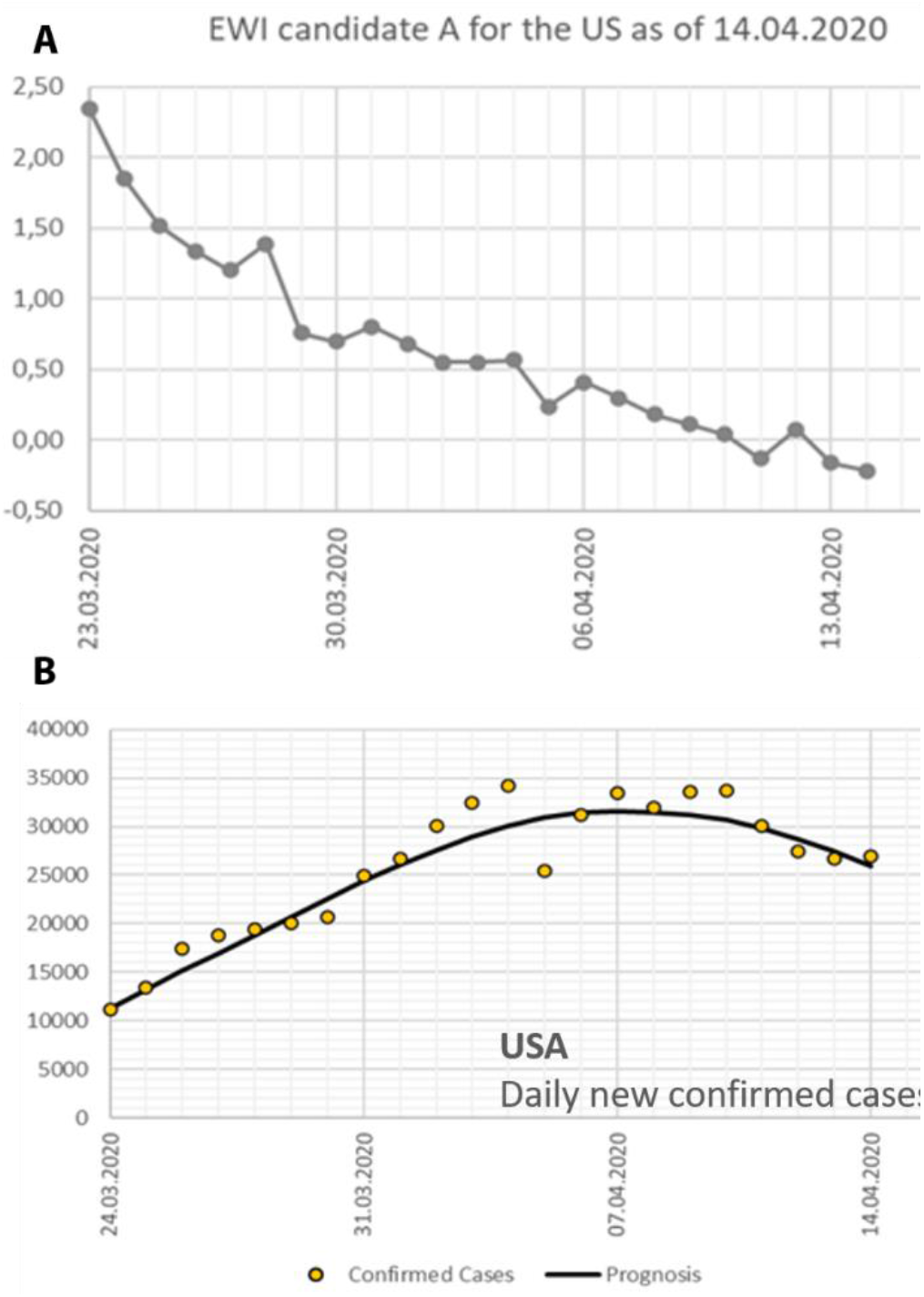
(A) EWI candidate A for the US 24.03.2020 – 14.04.2020. EWI candidate is a measure of whether the spread of the epidemic is currently accelerating (>0) or slowing down (<0). (B) Daily confirmed cases in the US according to worldometers.info

### Long-term prognosis

Based on the EWIs, coming out of the feature selection process, we modeled the underlying trends driving the dynamics of the epidemics. Such a model can be used for ongoing prediction and for an early sign for the risk of another outbreak. Preliminary results are as follows (see Figure 4).

**Figure 4:**
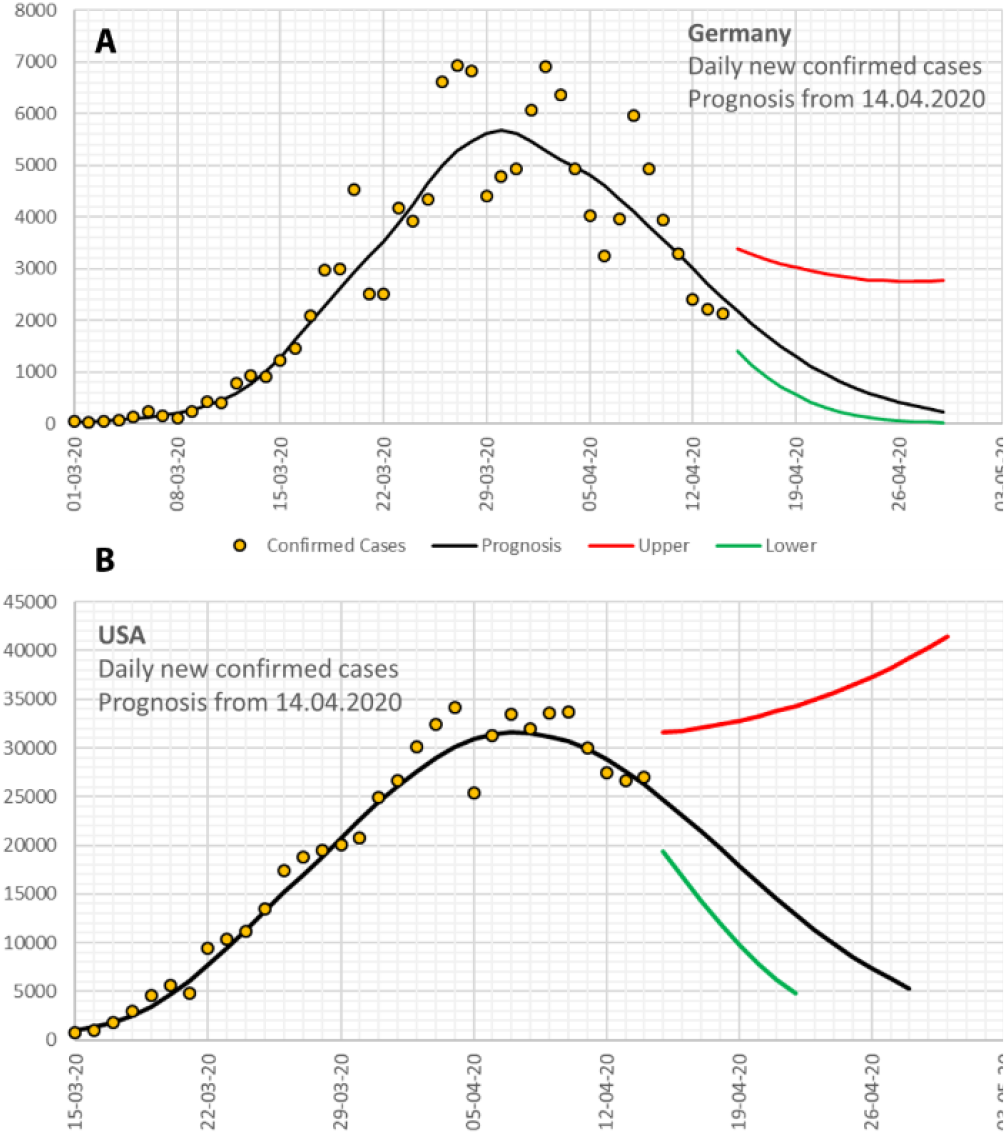
Epidemic projections along with its uncertainty according to the top-down model developed on 14.04.2020 for (A) Germany and (B) the USA. The dotted points show actual values for daily confirmed cases

### Germany

The first results confirm that the model used so far is accurate at least for a period of 7 to 10 days (Fig. 4A). For a longer time span, each prognosis must make assumptions e.g., on the underlying containment measures. Calculations show significant differences in the dynamics of the epidemics. In South Korea, the outbreak slowed down within two weeks, whereas in European countries, everything takes much longer. There is also a risk that in other countries, containment may fail completely.

### United States of America

This prognosis is valid only if the same level of containment measures is enforced in the whole country (Fig. 4B). Note that already small changes in containment measures may have a considerable impact on the prognosis, especially concerning the peak (number of infected cases can easily be 2-10 times higher). Here our prognosis can only be used to demonstrate what is to be expected if conditions of containment will not change, i.e., a best-case scenario.

### Short-term prognosis

Despite the limited utility of short-term transient trend for the number of newly infected cases and new deaths, we see merit in these such numbers for a two-fold reason.

Firstly, such just-in-time forecasts are the need of the hour, on which government agencies and institutions rely. It serves as a real-time tracking metric, describing what the current status is^16^. Just as the short-term forecast itself, decisions and judgments made from interpreting these numbers are equally transient.

Secondly, from a psychological and sociological perspective, people in the society need to base their daily outlook on data-driven evidence. Such prognosis numbers, if proven to be reliable as per our proposed framework, allow communities to be prepared and partake in compliance activities. Just as one consumes the weather forecast to be well equipped when leaving the house, a short-term prognosis with accuracy as good as the weather forecast aims to fulfill the same objective.

### Towards the development of a web-based platform

We have implemented these short-term prognoses in an online platform corona.quodata.de (see Figure 5). As of April 16, 2020, such prognoses can be accessed for 401 different districts and cities across Germany and 23 different countries on the portal. More national and sub-national entities will be added in the coming days. The table comprises miniature plots for (a) cumulative cases in the last 21 days as a line plot and (b) daily new confirmed cases as a bar plot. It reports the average doubling time in days for the number of confirmed cases based on a 7-day average relative growth rate. As per April 16, 2020, the average doubling time for Germany was 28 days. But if individual districts and counties are assessed, we see doubling times being as low as four days and as high as 70 days. These differences reflect the following facts (1) the spread of the pandemic varies considerably from region to region and (2) the decline in new infections is faster in some regions than in others.

**Figure 5:**
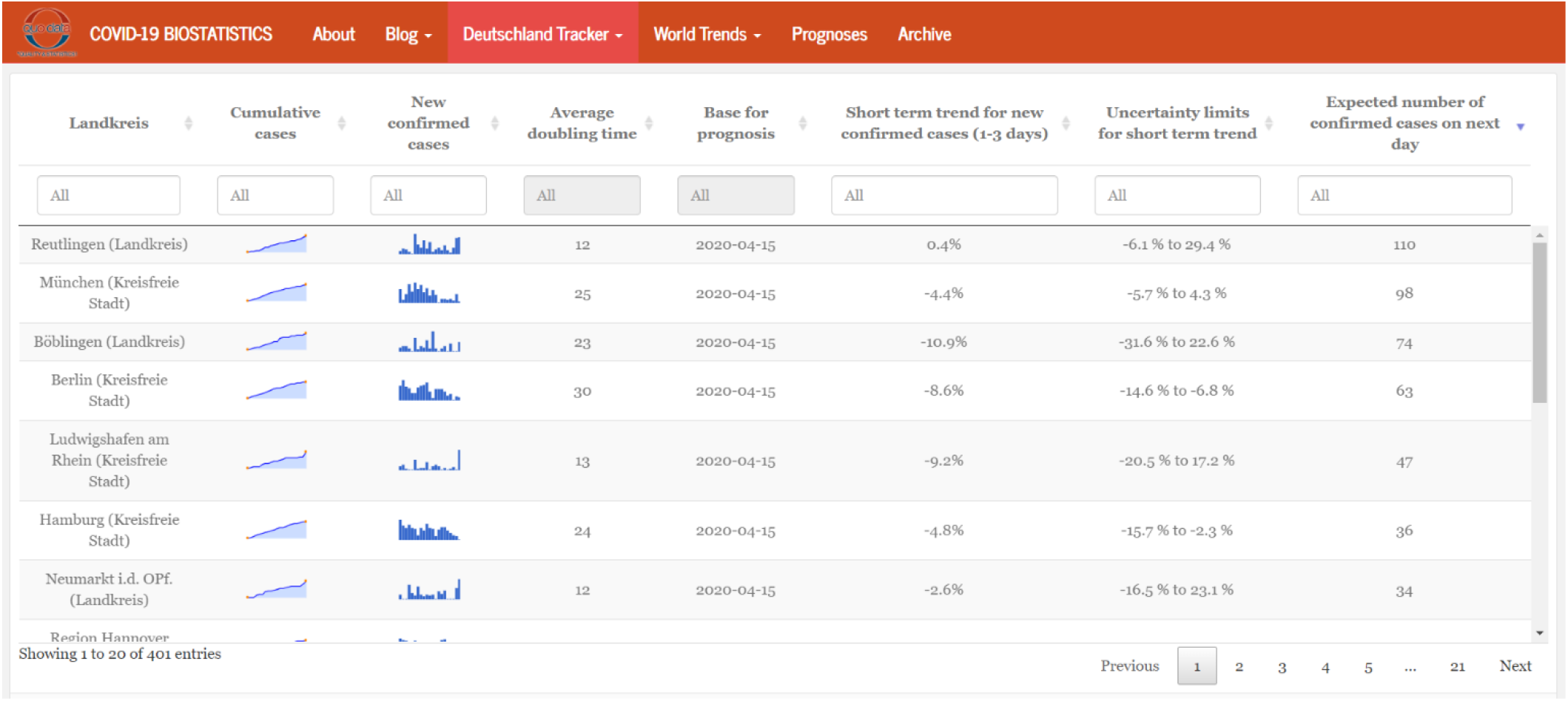
Screenshot for web-based portal on corona.quodata.de, with the short-term prognosis implemented for different cities and districts for Germany.

The table also reports “short term trend for confirmed cases”. This is essentially an interim prognosis obtained from the empirical top-down approach, which is the estimated percent change in the absolute daily number of cases for the next one to three days. The corresponding 80% uncertainty intervals are provided in the adjacent column. Lastly, the expected new cases are reported in the last column, calculated using the short-term trend. A key to tackling the epidemic growth or avoiding a resurgence is ease lockdown measures in cities and counties having low risks of a (renewed) outbreak, by monitoring the short-term prognosis, where business can resume partially with relaxed lockdown measures. And at the other end, keep strict guidelines and rules in place for cities and counties doubling their cases at a faster rate. It can be argued that such a coordinated shutdown-resume cycle for small geographical entities can limit the long-term socio-economic damage caused by a nationwide permanent lockdown.

We believe that the availability of such a web-based tool during such a challenging public health crisis not only keeps misinformation at bay (stemming out of daily change in case numbers) but provides a facile interpretation of the pace epidemic progression. To the best of our knowledge, we have not come across an implementation of an automatic short-term epidemic forecast system, which utilizes early warning indicators to provide quantitative insights into the underlying trend.

Although the built models have undergone several iterations of critical appraisals, in all likelihood, due to changing ‘ground situations’ in the coming days, the prediction of new cases can be incorrect for some national or subnational regions. The reader is advised to use the web-based portal and epidemic projections herein for guidance only, whilst being aware of radical new scenarios which may arise, throwing off the model in the current form.

## CONCULSIONS AND OUTLOOK

In summary, the results for EWIs show a remarkable correlation between case numbers and real-time change in the effective reproduction number. EWI for South Korea unequivocally describes the (a) peak of epidemic, days in advance, (b) predicts local outbreaks seen in the existence of multiple waves, and (c) pronounced effect of quick and decisive actions of the government. For Germany, EWI reflects a gradual decrease in epidemic potential in the ensuing days. Lastly, for the US, EWI presents a picture of a creeping decrease in epidemic potential, with fewer fluctuations compared to Germany.

Thus, EWIs reveal the course for different countries and reflect the effect of actions taken by the regions’ respective governments. We see potential in such EWIs, which we would like to exploit further. It should be noted that the EWI discussed here is one of the many indicators which show potential in modeling a realistic epidemic scenario. In our additional studies, we will rely heavily on neural networks to parse more and more region-specific EWIs.

We hope to integrate such an intelligent system in the online platform to evaluate data quality, uncertainties associated with estimated parameters, uncertainties associated with the model, track changes for a battery of EWIs, all in real-time. Such a dynamic, top-down modeling apapproachiteratively updates the prior distributions of parameterizationsand initializations and tunes the prognosis and its uncertainty. Altogether, all efforts focused to better equip individuals and institutions, to make informed decisionsand get insights into installed control policies.

## Data Availability

Data used in the manuscript has been sourced from publicly available databases as cited within the manuscript at relevant locations.

## ASSOCIATED CONTENT

Additional content can be found on https://corona.quodata.de

## REFERENCES

1. Biggerstaff, M. et al. Results from the centers for disease control and prevention’s predict the 2013--2014 Influenza Season Challenge. BMC Infect. Dis. 16, 357 (2016).

2. Chretien, J.-P., George, D., Shaman, J., Chitale, R. A. & McKenzie, F. E. Influenza forecasting in human populations: a scoping review. PLoS One 9, (2014).

3. Kukkonen, J. et al. A review of operational, regional-scale, chemical weather forecasting models in Europe. Atmos. Chem. Phys. 12, 1–87 (2012).

4. Morgan, O. How decision makers can use quantitative approaches to guide outbreak responses. Philos. Trans. R. Soc. B 374, 20180365 (2019).

5. Fauci, A. S., Lane, H. C. & Redfield, R. R. Covid-19—navigating the uncharted. (2020).

6. Pastor-Satorras, R., Castellano, C., Van Mieghem, P. & Vespignani, A. Epidemic processes in complex networks. Rev. Mod. Phys. 87, 925 (2015).

7. Bai, Z. et al. The Rapid Assessment and Early Warning Models for COVID-19. Virol. Sin. 1 (2020).

8. Enserink, M. & Kupferschmidt, K. Mathematics of life and death: How disease models shape national shutdowns and other pandemic policies. Sci. Mag. (2020).

9. Ferguson, N. et al. Report 9: Impact of non-pharmaceutical interventions (NPIs) to reduce COVID19 mortality and healthcare demand. (2020).

10. Lourenço, J. et al. Fundamental principles of epidemic spread highlight the immediate need for large-scale serological surveys to assess the stage of the SARS-CoV-2 epidemic. medRxiv (2020).

11. (IHME), T. I. for H. M. and E. COVID-19 Projections. Available at: https://covid19.healthdata.org/united-states-of-america. x(Accessed: 16th April 2020)

12. Covid, I., Murray, C. J. L. & others. Forecasting COVID-19 impact on hospital bed-days, ICU-days, ventilator-days and deaths by US state in the next 4 months. medRxiv (2020).

13. Viboud, C., Simonsen, L. & Chowell, G. A generalized-growth model to characterize the early ascending phase of infectious disease outbreaks. Epidemics 15, 27–37 (2016).

14. Dong, E., Du, H. & Gardner, L. An interactive web-based dashboard to track COVID-19 in real time. Lancet Infect. Dis. (2020).

15. worldometers.info. (2020). Available at: https://www.worldometers.info/. (Accessed: 13th April 2020)

16. Bettencourt, L. M. A. & Ribeiro, R. M. Real time bayesian estimation of the epidemic potential of emerging infectious diseases. PLoS One 3, (2008)

